# Sustained negative mental health outcomes among healthcare workers over the first year of the COVID-19 pandemic: a prospective cohort study

**DOI:** 10.1101/2021.11.21.21266594

**Authors:** Roberto Mediavilla, Eduardo Fernández-Jiménez, Irene Martinez-Morata, Fabiola Jaramillo, Jorge Andreo-Jover, Inés Morán-Sánchez, Franco Mascayano, Berta Moreno-Küstner, Sergio Minué, José Luis Ayuso-Mateos, Richard A. Bryant, María-Fe Bravo-Ortiz, Gonzalo Martínez-Alés, the COVID-19 HEalth caRe wOrkErS – Spain (HEROES-SPA) Group

## Abstract

**Objective:** To characterize the evolution of healthcare workers’ mental health status over the 1-year period following the initial COVID-19 pandemic outbreak and to examine baseline characteristics associated with resolution or persistence of mental health problems over time.

**Methods:** We conducted an 8-month follow-up cohort study. Eligible participants were healthcare workers working in Spain. Baseline data were collected during the initial pandemic outbreak. Survey-based self-reported measures included COVID-19-related exposures, sociodemographic characteristics, and three mental health outcomes (psychological distress, depression symptoms, and posttraumatic stress disorder symptoms). We examined three longitudinal trajectories in mental health outcomes between baseline and follow-up assessments (namely *asymptomatic/stable, recovering*, and *persistently symptomatic/worsening*).

**Results:** We recruited 1,807 participants. Between baseline and follow-up assessments, the proportion of respondents screening positive for psychological distress and probable depression decreased, respectively, from 74% to 56% and from 28% to 21%. Two-thirds remained asymptomatic/stable in terms of depression symptoms and 56% remained symptomatic or worsened over time in terms of psychological distress.

**Conclusions:** Poor mental health outcomes among healthcare workers persisted over time. Occupational programs and mental health strategies should be put in place.

---

The COVID-19 pandemic outbreak has had substantial mental health impact on healthcare workers (HCWs), largely due to increases in healthcare capacity requirements driving job redeployments and extended working hours in combination with very high risk of contagion and death. Estimates from cross-sectional studies conducted during the initial pandemic outbreak suggest that between 25 and 50% of HCWs may have experienced clinically significant symptoms of anxiety or depression (1–7) and posttraumatic stress (1,3,7).

The extent to which the deleterious mental health effects brought about by the initial pandemic outbreak may have subsequently led to negative mid- and long-term mental health outcomes among HCWs. However, these remain largely unexplored, despite important public health and clinical implications (8). Initially, it seemed plausible that a large proportion of the burden of mental health symptoms initially reported by HCWs would eventually resolve, either following cessation of exposure to the acute stressor or after adequate targeted interventions (e.g., self-care and low-intensity psychotherapeutic interventions). Nevertheless, because the initial pandemic outbreak has been followed by a series of ongoing subsequent pandemic waves that continue to strain health systems across the globe, there is a generalized concern that a substantial proportion of HCWs may be experiencing persistent mental health problems. According to the World Health Organization, reducing the long-term mental health impact of the pandemic on HCWs is considered a major clinical and public health priority. Estimating the mid- and long-term mental health impact of the COVID-19 pandemic among HCWs is paramount for occupational and mental healthcare planning purposes. In addition, identifying risk and protective factors for persistence of clinically significant mental health burden can help improve implementation of evidence-based detection and treatment strategies. Notwithstanding, evidence examining prevalence and predictors of persistence of mental health symptoms among HCWs from longitudinal cohort studies is scarce (9–12).

Here we used a large sample of HCWs during the one-year period following the initial pandemic outbreak in one of the largest COVID-19 hotspots globally to (1) characterize the evolution of HCWs’ mental health status over the year following the initial pandemic outbreak, and (2) examine baseline sociodemographic and clinical characteristics associated with resolution or persistence of mental health problems over time.

## Methods

### Study design, setting, and participants

We conducted a prospective cohort study in Spain as part of an ongoing longitudinal multi-national study (https://mentalnet.cl/en/home/). We collected data through an online survey at two timepoints. Baseline assessments were performed from April 24^th^ to June 22^nd^, 2020 (during the initial pandemic outbreak in Spain). Follow-up assessments took place between January 26^th^ and March 8^th^, 2021, which in Spain coincided with the third pandemic wave and with administration of COVID-19 vaccines for the majority of HCWs.

The study participants were HCWs aged 18 years and older, recruited from different outpatient and inpatient healthcare facilities, with clinical and non-clinical duties, and not necessarily involved in the direct care of COVID-19 patients. The sampling strategy was as follows. During both the baseline and follow-up assessments, key stakeholders (e.g., hospital managers, heads of worker unions) from healthcare facilities located in the study regions (Andalusia, Madrid, and Murcia) forwarded the survey link to all HCWs. Participants were also asked to forward the survey to peers in order to enhance response rates. In addition, during the follow-up period, we sent email or telephone survey reminders to baseline participants. Baseline assessments are described elsewhere (13). In this manuscript, we focus on participants who were assessed at follow-up only and on participants assessed at follow-up who had been assessed also during baseline procedures. We hereafter refer to these two subgroups as *partial* and *full respondents*, respectively.

All procedures contributing to this work comply with the Helsinki Declaration of 1975, as revised in 2013. It received approval from the Hospital La Paz Ethics Committee (Madrid, Spain) and was ratified by the local ethics committees from the participating sites.

### Variables

Baseline assessments included the following COVID-19-related exposures: direct involvement in the care of COVID-19 patients (yes, no), adequate access to personal protective equipment, fear of getting infected, and fear of infecting loved ones (all rated from 0 to 3).

Both baseline and follow-up assessments included the following sociodemographic and mental health outcome variables.

### Sociodemographic variables

Age in years, gender (male, female), educational level (primary, secondary, or university studies), and type of job. We collapsed job types into the following categories: physicians, nurses, health technicians (e.g., nurse, X-ray, and laboratory technicians), ancillary workers (e.g., security staff, drivers, administrative staff, and cleaning staff), other HCWs (e.g., clinical psychologists, physiotherapists, and biologists), and residential support workers (e.g., from mental health assisted living facilities, nursing homes).

### Mental health outcomes

Psychological stress, as measured by the Spanish version of the 12-item General Health Questionnaire (14); and probable depression symptoms, as measured by the Spanish version of the 9-item Patient Health Questionnaire (PHQ-9) (15). We used widely accepted thresholds for detecting people screening positive for psychological distress (GHQ-12 higher than 2 points) (16,17) and for depression (PHQ-9 score higher than 9 points) (18). In addition, follow-up assessments also included posttraumatic stress disorder (PTSD) symptoms, as measured by the Spanish 5-item version of the Primary Care PTSD Screen for DSM-5 (PC-PTSD-5), where a total score higher than 2 points suggests probable PTSD (19).

Cronbach’s alphas were 0.87 (95 percent CI: 0.86, 0.88) for the GHQ-12 total score; 0.89 (95 percent CI: 0.88, 0.89) for the PHQ-9 total score; and 0.70 (95 percent CI: 0.68, 0.72) for the PC-PTSD-5. To control for region-level cumulative COVID-19 incidence, we calculated region-specific 14-day cumulative incidence rates 2, 4, 6, and 8 weeks after the start of the follow-up period and, as rates were stable over time, classified regions as “high” or “low” incidence depending on whether average cumulative incidence over time points fell under or over 750 cases per 100,000 based on visual examination of region-specific cumulative incidence rates (**Supplementary Figures 1 and 2)**.

### Statistical analyses

First, we removed baseline respondents who provided informed consent but did not go on to initiate the survey (n = 95). We reported categorical variables as frequencies and valid percentages, and continuous and interval variables as either mean (standard deviation [SD]), or median (interquartile range [IQR]). Descriptive statistics were calculated separately for full and partial respondents.

Then, we used multivariable mixed-effects linear and binary logistic regression models to explore the associations between baseline variables, including sociodemographic characteristics (i.e., age, gender, and educational level) and COVID-19-related exposures (i.e., direct involvement in the care of COVID-19 patients, adequate access to personal protective equipment, fear of getting infected, and fear of infecting loved ones), and follow-up mental health outcomes (i.e., psychological distress, depression symptoms, and PTSD symptoms), defined both as continuous questionnaire scores and dichotomous variables. We conducted sensitivity analyses adjusted by baseline assessments of the follow-up outcome under consideration. We used baseline GHQ-12 score for the model where follow-up PTSD was the outcome, as we did not have estimates for the latter in the baseline assessment. The GHQ-12 score, an instrument that has good convergent validity with the PC-PTSD-5 and accurately detects PTSD in primary care settings (20).

Next, we used baseline and follow-up mental health outcomes to categorize respondents into three mental health trajectories, separately for psychological distress and for depression, according to whether they screened negative at baseline and follow-up (*asymptomatic stable*), positive at baseline and negative at follow-up (*recovering*), or positive or negative at baseline and positive at follow-up (*persistently symptomatic/worsening*). For instance, if a respondent screened negative in the GHQ-12 and positive in the PHQ-9 at baseline, and subsequently screened positive in the GHQ-12 and negative in the PHQ-9 at follow-up, they would belong to the *persistently symptomatic/worsening* trajectory for psychological distress and to the *recovering* trajectory for depression. We selected these trajectory categories because of their potential implications for clinical practice.

Finally, we explored the association between baseline exposures and longitudinal psychological distress and depression trajectory membership, using multinomial regression models where *asymptomatic stable* was considered the reference category. Baseline exposures were age group, gender, educational level, direct involvement in the care of COVID-19 patients, adequate access to personal protective equipment (adequate vs. inadequate), fear of getting infected (not or slightly afraid vs. considerably or extremely afraid), and fear of infecting loved ones (not or slightly afraid vs. considerably or extremely afraid).

All models were adjusted by covariates to control for confounding based on prior causal knowledge, using direct acyclic graphs and backdoor criteria (21). Region-level cumulative incidence of COVID-19 was entered in all models as a fixed factor. We did not impute missing data. All analyses were performed using packages *dplyr, gtsummary, flextable, ggplot2, psych, multinom* of R Studio for Mac (Version 1.2.5042).

## Results

Of 1,807 respondents who answered the survey at follow-up (between January 26^th^-March 25^th^, 2021), 1,471 (81.4%) completed the entire survey, with a median response time of 21 minutes. Most missing data pertained to the last section of the questionnaire, suggesting that data missingness was driven by survey extension and hence largely random. Respondents who did and did not complete the survey were comparable in terms of mean age (42 vs. 40 years, respectively) and gender distribution (78% vs. 74% female, respectively). Response rates were estimated across facilities and job types and ranged from 2.7% to 100% (see **Supplementary Tables 1 and 2**).

There were 1,058 (59%) partial respondents (i.e., assessed only at follow-up) and 749 (41%) full respondents (i.e., assessed at both baseline and follow-up). Of note, this indicates that we retained 32% of the 2,370 original baseline respondents for follow-up assessments (see **Figure 1**). Sociodemographic characteristics of follow-up respondents, overall and divided into full and partial respondents, are shown in **Table 1**. In short, full respondents were more frequently female and more likely to have completed university studies than partial respondents. Also, while most full respondents were physicians or nurses, partial respondents included a larger proportion of residential support workers.

**Figure 1.**
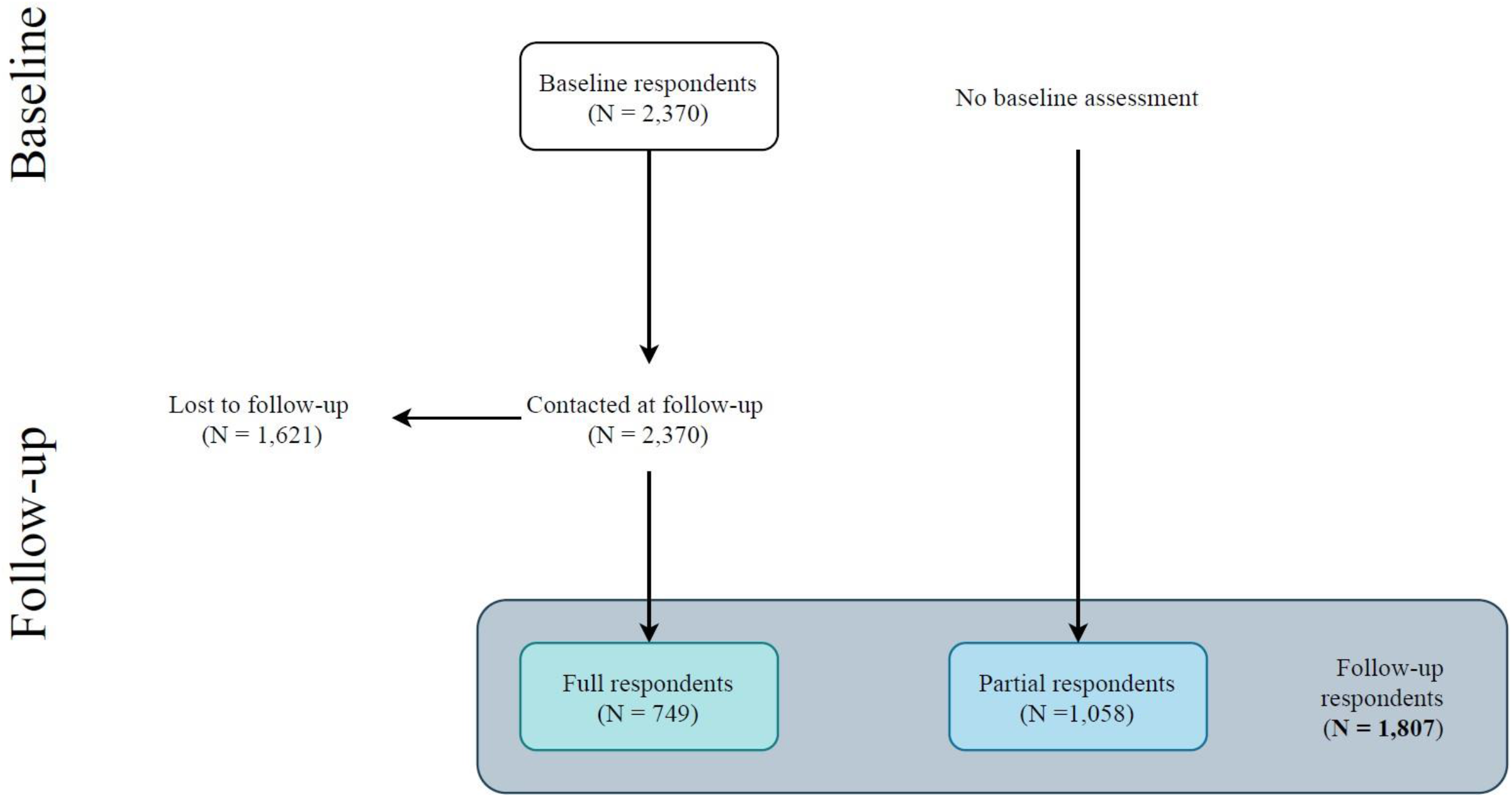
Flowchart of the participants. Follow-up respondents (N = 1,807) include participants who completed both baseline and follow-up assessments (i.e., *full respondents*) and participants who completed the follow-up assessment only (i.e., *partial respondents*). [The COVID-19 HEalth caRe wOrkErS (HEROES) Study, Spain, 2021]

**Table 1.** Characteristics of the participants who underwent baseline assessment (*full respondents*) and who did not (*partial respondents)* as measured at follow-up. [The COVID-19 HEalth caRe wOrkErS (HEROES) Study, Spain, 2021]

|  | All,<br>N = 1,807 | Baseline assessment |  |
| --- | --- | --- | --- |
|  |  | No (partial respondents),<br>N = 1,058 | Yes (full respondents),<br>N = 749 |
| Age (in years), *M* (SD) | 42 (11) | 42 (12) | 42 (11) |
| Missing | 90 | 46 | 44 |
| Gender, *n* (%) |  |  |  |
| Male | 412 (23) | 273 (26) | 139 (19) |
| Female | 1,368 (77) | 774 (74) | 594 (81) |
| Missing | 27 | 11 | 16 |
| Educational level |  |  |  |
| Primary studies | 18 (1.0) | 10 (1.0) | 8 (1.1) |
| Secondary studies | 397 (22) | 311 (30) | 86 (12) |
| University studies | 1,355 (77) | 720 (69) | 635 (87) |
| Missing | 37 | 17 | 20 |
| Type of job |  |  |  |
| Physicians | 419 (25) | 169 (17) | 250 (36) |
| Nurses | 312 (18) | 107 (11) | 205 (30) |
| Health technicians <sup>a</sup> | 86 (5) | 41 (4) | 45 (6) |
| Other HCWs <sup>b</sup> | 268 (16) | 188 (19) | 80 (12) |
| Ancillary workers <sup>c</sup> | 157 (9.3) | 119 (12) | 38 (5) |
| Residential support workers | 367 (22) | 312 (31) | 55 (8) |
| Other | 80 (5) | 59 (6) | 21 (3) |
| Missing | 118 | 63 | 55 |

*Note*
All percentages are valid percentages
HCWs = healthcare workers
<sup>a</sup> Health technicians include nurse, X-ray, or laboratory technicians, among others
<sup>b</sup> Other HCWs include clinical psychologists, physiotherapists, or biologists, among others
<sup>c</sup> Ancillary workers include security staff, drivers, administrative staff, or cleaning staff, among others

Overall, 56% of follow-up respondents screened positive for psychological distress, 21% for probable depression, and 51% for PTSD. Psychological distress, probable depression, and PTSD were more frequent among younger and female respondents, and respondents with higher educational levels – with substantial heterogeneity across specific job types (see **Supplementary Table 3**). Notably, follow-up mental health outcomes were comparable between full and partial respondents, with similar mean (SD) GHQ-12 score (3.9 [3.5] vs. 3.8 [3.4], respectively), mean (SD) PHQ-9 score (6.3 [5.1] vs. 6.4 [3.4], respectively), proportion of respondents screening positive for psychological distress (56% vs. 55%, respectively), and proportion of respondents screening positive for probable depression (21% vs. 21%, respectively).

Comparisons between baseline and follow-up mental health outcomes among full respondents are shown in **Table 2**. The proportion of respondents screening positive for psychological distress and probable depression decreased, respectively, from 74% to 56% and from 28% to 21%. **Figure 2** shows the distribution of trajectories of depression symptoms and psychological distress over time, overall and across baseline covariates. Trajectories show that, in terms of depression symptoms, 66% respondents remained asymptomatic/stable, 15% recovered, and 19% remained symptomatic or worsened over time. In terms of psychological distress, 18% respondents remained asymptomatic/stable, 26% recovered, and 56% remained symptomatic or worsened over time. The distribution of trajectories was heterogeneous across baseline covariates.

**Table 2.**
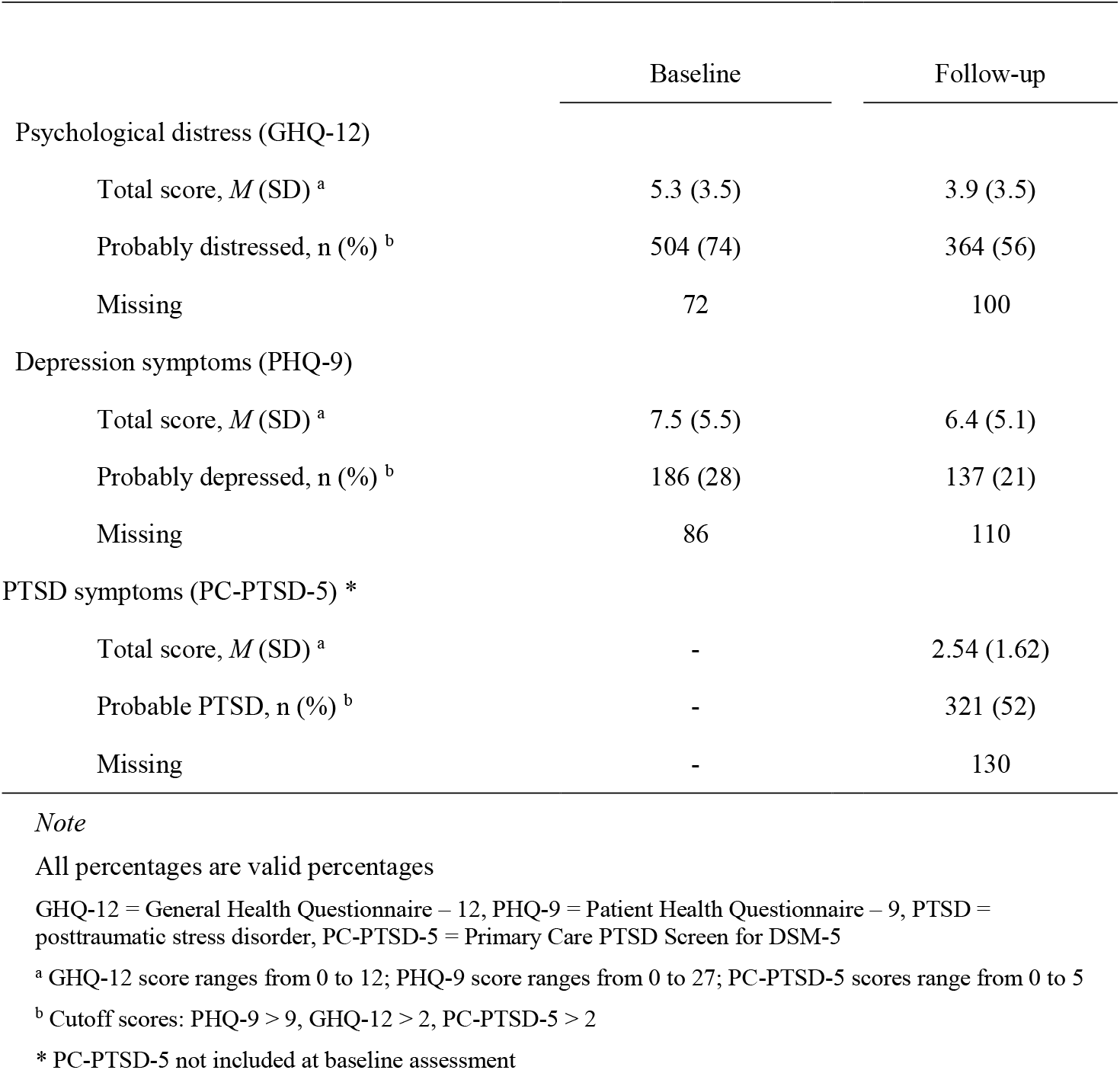
Mental health outcomes among *full respondents* (N = 749) at baseline and follow-up. [The COVID-19 HEalth caRe wOrkErS (HEROES) Study, Spain, 2021]

**Figure 2.**
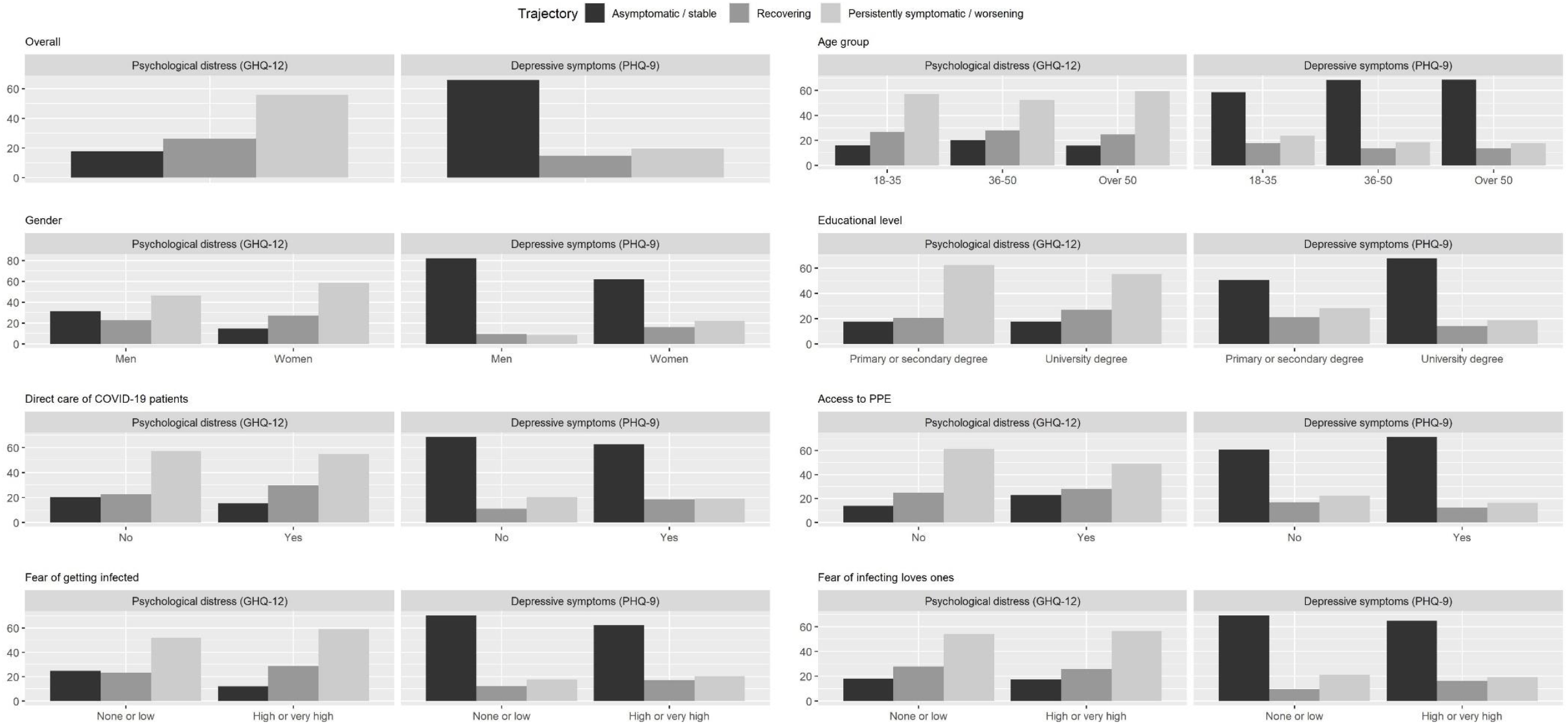
Mental health outcome trajectories of psychological distress and depression symptoms stratified by relevant variables. Trajectories include people with positive screening at baseline and negative screening at follow-up (*recovering*), people with positive or negative screening at baseline and positive screening at follow-up (*persistently symptomatic/worsening*) and people with negative screening at baseline and follow-up (*asymptomatic stable*). [The COVID-19 HEalth caRe wOrkErS (HEROES) Study, Spain, 2021]

**Table 3** shows crude and adjusted estimates of the association between baseline exposures and follow-up mental health outcome scores among full respondents. Overall, women had higher scores (i.e., worse mental health) than men, and job-related factors such as direct involvement in the care of COVID-19, inadequate access to protective equipment, or fear of infecting oneself or loved ones were associated with higher negative mental health outcome scores - especially for PTSD, and with higher odds of testing positive for psychological distress, probable depression, and PTSD (see **Supplementary Table 4**). We repeated all models including further adjustment by baseline mental health outcome scores: results did not change substantially (see **Supplementary Tables 5 and 6**).

**Table 3.** Association between participants’ sociodemographic characteristics and COVID-19-related exposures, measured at baseline, and mental health outcomes’ total scores (psychological distress, depressive symptoms, and posttraumatic stress disorder symptoms), measured at follow-up (8 months). [The COVID-19 HEalth caRe wOrkErS (HEROES) Study, Spain, 2021]

|  | Psychological distress (GHQ-12) |  |  |  | Depression symptoms (PHQ-9) |  |  |  | PTSD symptoms (PC-PTSD-5) |  |  |  |
| --- | --- | --- | --- | --- | --- | --- | --- | --- | --- | --- | --- | --- |
|  | Unadjusted |  | Adjusted |  | Unadjusted |  | Adjusted |  | Unadjusted |  | Adjusted |  |
|  | B | 95 percent CI | B | 95 percent CI | B | 95 percent CI | B | 95 percent CI | B | 95 percent CI | B | 95 percent CI |
| Age in years <sup>a</sup> | 0.00 | (-0.03, 0.02) | 0.00 | (-0.02, 0.03) | -0.04 | (-0.08, 0) | -0.03 | (-0.07, 0.01) | -0.02 | (-0.04, -0.01) | -0.02 | (-0.03, -0.01) |
| Female gender <sup>b</sup> | 1.05 | (0.32, 1.77) | 1.07 | (0.33, 1.8) | 2.14 | (1.1, 3.18) | 2.06 | (1, 3.11) | 0.68 | (0.34, 1.02) | 0.64 | (0.3, 0.98) |
| Educational level <sup>c</sup> | -0.22 | (-0.57, 0.12) | -0.17 | (-0.52, 0.18) | -0.54 | (-1.02, -0.05) | -0.51 | (-1, -0.02) | -0.16 | (-0.32, 0) | -0.18 | (-0.34, -0.02) |
| Frontline position <sup>c</sup> | 0.13 | (-0.42, 0.69) | -0.04 | (-0.64, 0.56) | 0.08 | (-0.72, 0.89) | -0.51 | (-1.37, 0.35) | 0.46 | (0.2, 0.73) | 0.42 | (0.13, 0.7) |
| Adequate access to PPE <sup>c</sup> | -0.53 | (-0.86, -0.2) | -0.47 | (-0.81, -0.14) | -1.03 | (-1.5, -0.56) | -0.86 | (-1.34, -0.39) | -0.37 | (-0.52, -0.21) | -0.32 | (-0.48, -0.16) |
| Fear of getting infected <sup>c</sup> | 0.25 | (-0.13, 0.63) | 0.19 | (-0.19, 0.58) | 0.42 | (-0.12, 0.96) | 0.32 | (-0.24, 0.87) | 0.46 | (0.28, 0.64) | 0.44 | (0.26, 0.62) |
| Fear of infecting loved ones <sup>c</sup> | 0.32 | (-0.02, 0.66) | 0.35 | (0, 0.69) | 0.66 | (0.17, 1.16) | 0.73 | (0.24, 1.23) | 0.50 | (0.34, 0.65) | 0.50 | (0.34, 0.65) |

*Note*
GHQ-12 = General Health Questionnaire – 12 items, PHQ-9 = Patient Health Questionnaire – 9 items, PC-PTSD-5 = Primary Care PTSD Screen for DSM-5, B = beta, CI = confidence interval, PPE = personal protective equipment
<sup>a</sup> Adjusted for gender
<sup>b</sup> Adjusted for age
<sup>c</sup> Adjusted for age, gender, and region-level 14-day COVID-19 cumulative incidence (fixed factor)

**Table 4** shows crude and adjusted associations between baseline exposures and trajectory membership for psychological distress and probable depression. Women showed higher symptom variability over time than men, as indicated by women’s higher odds of belonging to both the *recovering* and the *persistently symptomatic/worsening* categories for both psychological distress and probable depression. In terms of psychological distress, reporting inadequate access to personal protective equipment was associated with *persistently symptomatic/worsening* category membership. In terms of probable depression, fear of infecting loved ones was associated with *recovering* category membership.

**Table 4.** Association between participants’ sociodemographic characteristics and COVID-19-related exposures, measured at baseline, and the probability of belonging to the trajectories *recovering* or *persistently symptomatic/worsening* (versus *asymptomatic stable*) at follow-up. [The COVID-19 HEalth caRe wOrkErS (HEROES) Study, Spain, 2021]

|  | Psychological distress (GHQ-12) |  |  |  |  |  |  |  | Depression symptoms (PHQ-9) |  |  |  |  |  |  |  |
| --- | --- | --- | --- | --- | --- | --- | --- | --- | --- | --- | --- | --- | --- | --- | --- | --- |
|  | Recovering |  |  |  | Persistently symptomatic / worsening |  |  |  | Recovering |  |  |  | Persistently symptomatic / worsening |  |  |  |
|  | Unadjusted |  | Adjusted |  | Unadjusted |  | Adjusted |  | Unadjusted |  | Adjusted |  | Unadjusted |  | Adjusted |  |
|  | OR | 95 percent CI | OR | 95 percent CI | OR | 95 percent CI | OR | 95 percent CI | OR | 95 percent CI | OR | 95 percent CI | OR | 95 percent CI | OR | 95 percent CI |
| 18-35 years old [ref: > 50] <sup>a</sup> | 1.06 | (0.53, 2.16) | 1.00 | (0.49, 2.04) | 0.94 | (0.5, 1.76) | 0.88 | (0.47, 1.66) | 1.53 | (0.81, 2.91) | 1.46 | (0.76, 2.77) | 1.56 | (0.88, 2.77) | 1.46 | (0.82, 2.62) |
| 35-50 years old [ref: > 50] <sup>a</sup> | 0.89 | (0.46, 1.7) | 0.83 | (0.43, 1.61) | 0.69 | (0.39, 1.23) | 0.64 | (0.36, 1.16) | 1.00 | (0.54, 1.87) | 0.96 | (0.51, 1.81) | 1.04 | (0.59, 1.81) | 0.99 | (0.56, 1.74) |
| Female gender [ref: male] <sup>b</sup> | 2.54 | (1.38, 4.68) | 2.54 | (1.38, 4.68) | 2.67 | (1.58, 4.5) | 2.67 | (1.58, 4.5) | 2.24 | (1.1, 4.59) | 2.45 | (1.16, 5.2) | 3.47 | (1.66, 7.25) | 3.40 | (1.62, 7.12) |
| University studies [ref: primary/secondary] <sup>c</sup> | 1.31 | (0.57, 3.02) | 1.82 | (0.74, 4.44) | 0.87 | (0.43, 1.75) | 1.09 | (0.52, 2.27) | 0.49 | (0.25, 0.96) | 0.45 | (0.22, 0.91) | 0.49 | (0.27, 0.9) | 0.48 | (0.25, 0.92) |
| Frontline position [ref: no] <sup>c</sup> | 1.75 | (1.05, 2.91) | 1.14 | (0.65, 2.01) | 1.28 | (0.82, 2.02) | 0.90 | (0.54, 1.49) | 1.79 | (1.09, 2.95) | 1.43 | (0.83, 2.46) | 1.01 | (0.66, 1.56) | 0.80 | (0.49, 1.29) |
| Adequate access to PPE [ref: inadequate] <sup>c</sup> | 0.69 | (0.41, 1.15) | 0.80 | (0.47, 1.37) | 0.49 | (0.31, 0.78) | 0.57 | (0.35, 0.92) | 0.63 | (0.38, 1.04) | 0.71 | (0.43, 1.19) | 0.63 | (0.4, 0.98) | 0.72 | (0.45, 1.14) |
| Fear of getting infected [ref: low] <sup>c</sup> | 2.58 | (1.5, 4.45) | 2.22 | (1.25, 3.93) | 2.34 | (1.44, 3.81) | 2.07 | (1.24, 3.45) | 1.59 | (0.95, 2.65) | 1.42 | (0.83, 2.41) | 1.31 | (0.83, 2.07) | 1.23 | (0.76, 1.98) |
| Fear of infecting loved ones [ref: low] <sup>c</sup> | 0.97 | (0.52, 1.82) | 1.51 | (0.76, 2.98) | 1.09 | (0.62, 1.92) | 1.57 | (0.85, 2.91) | 1.75 | (0.87, 3.5) | 2.10 | (1.02, 4.32) | 0.97 | (0.57, 1.64) | 1.09 | (0.62, 1.92) |

*Note*
GHQ-12 = General Health Questionnaire – 12 items, PHQ-9 = Patient Health Questionnaire – 9 items, OR = odds ratio, CI = confidence interval, PPE = personal protective equipment
*Recovering*: positive screening at baseline and negative screening at follow-up; *persistently symptomatic/worsening*: positive or negative screening at baseline and positive screening at follow-up; *asymptomatic stable*: negative screening at baseline and follow-up
<sup>a</sup> Adjusted for gender
<sup>b</sup> Adjusted for age
<sup>c</sup> Adjusted for age, gender, and region-level 14-day COVID-19 cumulative incidence (fixed factor)

## Discussion

This study followed a cohort of HCWs from one of the earliest COVID-19 pandemic hotspots over the one-year period after the initial pandemic outbreak. There was marked heterogeneity across individuals in terms of variation in mental health outcomes over time. While we detected general reductions in psychological distress (from 74 to 56%) and depression symptoms (from 28 to 21%), the overall burden of poor mental health among HCWs remained substantial 8 months after the pandemic onset (56% screened positive for psychological distress, 21% for probable depression, and 51% for PTSD). Our analysis of mental health outcome trajectories revealed that psychological distress and depression symptoms persisted or worsened over time for 56% and 19% of respondents, respectively. We identified prospective associations between certain baseline characteristics, such as being female, reporting inadequate access to personal protective equipment, or being afraid of getting infected and of infecting loved ones, and follow-up psychological distress, depression symptoms, and PTSD symptoms. These results highlight the importance of adapting, implementing, and scaling-up evidence-based public, occupational, and mental health interventions for HCWs to prevent their mental health from further deteriorating during the ongoing pandemic and its aftermath.

Early cross-sectional studies from high-incidence COVID-19 areas, such as the Chinese region of Wuhan (22), Italy (3,5), or Spain (1), described the mental health toll taken by the pandemic on HCWs’ mental health, showing remarkable rates of psychological distress, anxiety, depression, and PTSD symptoms. In our study sample, estimates of the point prevalence of depression at baseline were similar than those found in Italy (25%) and Spain (28%), probably due to similar sample characteristics and study settings. Likewise, our baseline finding that three in four respondents were psychologically distressed is nearly identical to that of Lai and colleagues in Wuhan at the beginning of the pandemic (late January 2020). Additionally, a series of cross-sectional studies had already reported associations between HCWs’ characteristics, such as female gender, or inadequate access to personal protective equipment and negative mental health outcomes (i.e., anxiety or depression) (23–25). Our study found these associations to persist within a prospective cohort design, lowering the risk of potential reverse causation bias and hence greatly enhancing interpretability for decision-making.

Other prospective studies have sought to describe the evolution of HCWs’ negative mental health outcomes over time using a variety of outcomes and follow-up periods (9–12,25–28). Somewhat in contrast to our results, López Steinmez and colleagues reported a slight increase in psychological distress (from 40% to 46% point prevalence) between May and September, 2020, in a sample of 300 HCWs from Buenos Aires, Argentina. Differences in follow-up time probably accounts for this between-study difference, as they may have captured the early consequences of the initial pandemic outbreak while we conducted our assessments later, when renovated reasons for optimism (e.g., vaccine development and roll-out) had already started to emerge. Using a highly homogeneous sample of 200 nurses, Pinho and colleagues reported stable trends in depression and decreasing trends in anxiety between April and November, 2020. Differences in sample composition make their results hardly comparable to our’s. Other studies have either used much shorter follow-up periods (25,27,28) or reported outcomes not comparable to our’s, such as insomnia (26) or job stress (12). Our’s is the first prospective cohort analysis of risk factors for negative mental health outcomes among HCWs to adjust all associations of interest for potential confounding due to area-level COVID-19 cumulative incidence, in addition to adjustment for individual-level confounding. Notably, mounting evidence suggests higher rates of negative mental health outcomes among HCWs from regions with higher incidence (29).

To our knowledge, only one study has described the trajectories of mental health problems among HCWs during the COVID-19 pandemic (9). Using latent class modelling based on scores on three mental health outcomes (depression, anxiety, and PTSD symptoms) between May and September, 2020, they found four distinct trajectories which are remarkably similar to ours in terms of interpretation and prevalence within the study sample: 19% respondents belonged to their ‘recovered’ group (for 15% in our *recovering* group), 66% to their ‘resilient’ group (for 66% in our *asymptomatic stable* group), and 7% and 8%, respectively, to their ‘sub-chronic’ and ‘delayed’ groups (for 19% in our *symptomatic/worsening* group). These same trajectories have been identified is studies using latent growth mixture modelling across many different populations that have experienced adversity (30). Notably, this previous study did not assess psychological distress. Accordingly, our surprisingly high rates of persistence or worsening of psychological distress (56% of respondents) cannot be compared to other studies. While this result does not lend itself to easy interpretation until subsequent follow-up studies using the GHQ-12 emerge, it seems plausible that a substantial proportion of HCWs may potentially beneficiate from implementation of programs to lower psychological distress.

In addition to confirming associations previously reported in cross-sectional reports, our findings expand existing evidence in impactful ways for public health and clinical decision-making. First, by including a heterogeneous sample of HCWs with and without clinical duties, our study may serve to inform strategies aimed at non-clinical workers such as administrators or cleaners – largely overlooked in most studies examining mental health outcomes among HCWs during the COVID-19 pandemic. Second, our finding of scarce evidence of reliable baseline predictors of mental health outcome trajectories over time suggests that all HCWs should be offered easy-to-access mental health resources tailored to their needs (i.e., self-care and low-intensity psychotherapeutic interventions), regardless of profile in terms of sociodemographic characteristics and baseline clinical features.

Our study has limitations. First, we used a non-random sample that increases probability of some degree of collider bias and hinders transportability of study results across settings. Also, and in line with other multi-center studies (1,4), response rates varied significantly across sites and facilities, and the possibility of self-selection bias cannot be ruled out. Nevertheless, the baseline sociodemographic characteristics and mental health outcomes were however similar to another Spanish study with a larger and somewhat more representative sample of HCWs (1) and to other, similar European studies (2,3). Second, because of the use of observational data, effect estimates are potentially subject to some degree of residual confounding. Notably, substantial residual confounding is unlikely given that we included measures on all major individual- and region-level confounders and that estimates from crude and adjusted associations are roughly similar. Moreover, sensitivity analyses exploring differences between subsamples (e.g., full vs. partial respondents) and adjusting for baseline measurements of mental health outcomes obtained similar results, suggesting that our models were robust to different model specifications. Third, two thirds of baseline respondents were lost to follow-up. Dropout was independent from age, gender, and mental health outcomes at baseline, but people lost to follow were slightly more concerned about getting the virus and infecting their loved ones (data not shown). Fourth, limitations of self-reports for diagnostic screening are widely known (31). In the context of HCWs’ reactions to an initial pandemic outbreak, available diagnostic thresholds might have misclassified early, adaptive reactions to acute stressors as probable disorders (i.e., false positives). Notwithstanding, we used widely accepted screening instruments with good psychometric properties validated worldwide. Last, we calculated outcome trajectories based on information from two time points only. Future steps will include ascertainment of mental health outcomes in subsequent follow-up assessments and adoption of data-driven latent growth modelling approaches in addition to previously established categories based on clinical implications.

This is the first study to describe the trajectories of change of a large sample of HCWs from an early pandemic hotspot over a long follow-up period. Our results suggest preventative and restorative strategies at various levels (i.e., public, occupational, and specialized mental health), and outlines modifiable factors that might inform resource allocation, such as provision of protective equipment or being in direct care of COVID-19 patients. Further studies exploring the long-term impact of the pandemic among HCWs are warranted.

## Data Availability

All data produced in the present study are available upon reasonable request to the authors

