## Supplementary for "Sustained negative mental health outcomes among healthcare workers over the first year of the COVID-19 pandemic: a prospective cohort study"

Supplementary material

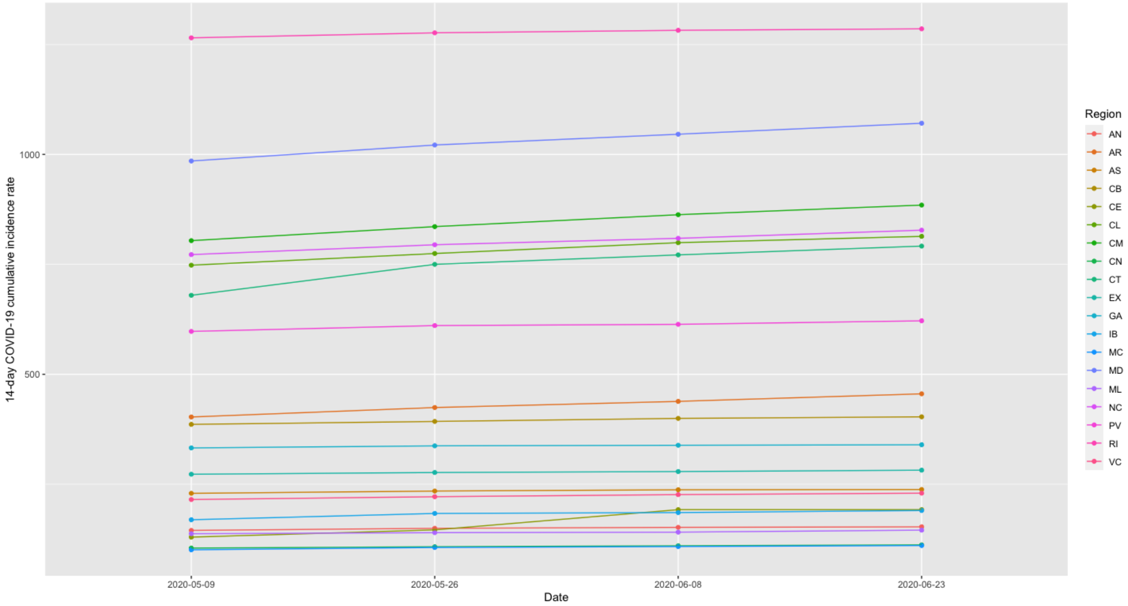


**Supplementary Figure 1**. 14-day cumulative incidence by region at 2, 4, 6 and 8 weeks after the study onset. AN = Andalucía, AR = Aragón, AS = Asturias, CB = Cantabria, CE = Ciudad Autónoma de Ceuta, CL = Castilla y León, CM = Castilla-La Mancha, CN = Canarias, CT = Cataluña, EX = Extremadura, GA = Galicia, IB = Islas Baleares, MC = Región de Murcia, ML = Ciudad Autónoma de Melilla, NC = Comunidad Foral de Navarra, PV = País Vasco, RI = La Rioja, VC = Comunidad Valenciana. [The COVID-19 HEalth caRe wOrkErS (HEROES) Study, Spain, 2021]

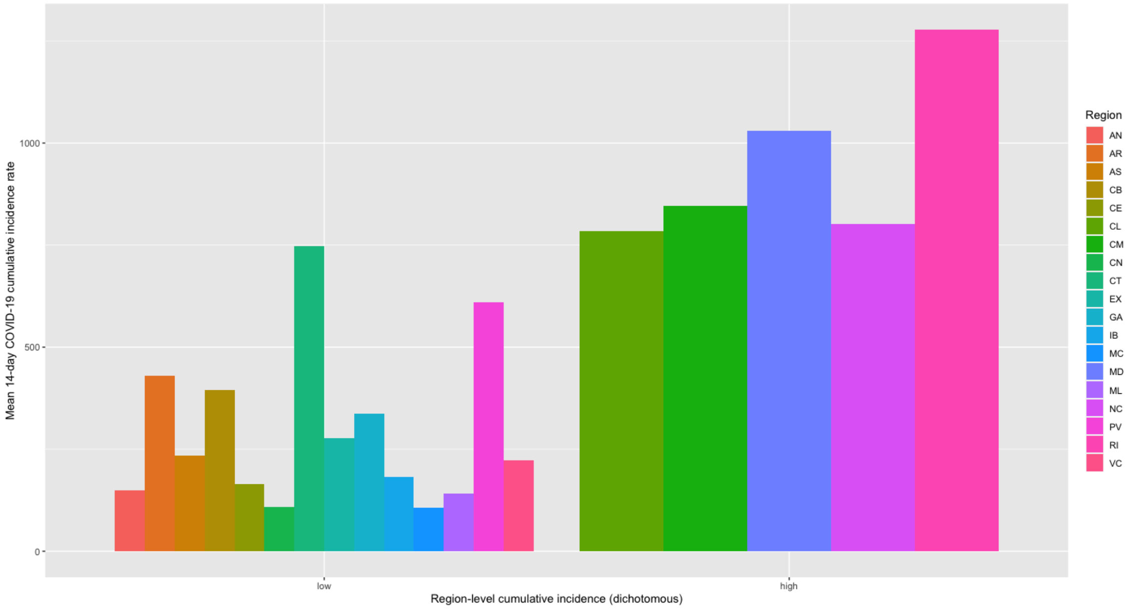


**Supplementary Figure 2**. 14-day cumulative incidence by region at 2, 4, 6 and 8 weeks after the study onset (bis). AN = Andalucía, AR = Aragón, AS = Asturias, CB = Cantabria, CE = Ciudad Autónoma de Ceuta, CL = Castilla y León, CM = Castilla-La Mancha, CN = Canarias, CT = Cataluña, EX = Extremadura, GA = Galicia, IB = Islas Baleares, MC = Región de Murcia, ML = Ciudad Autónoma de Melilla, NC = Comunidad Foral de Navarra, PV = País Vasco, RI = La Rioja, VC = Comunidad Valenciana. [The COVID-19 HEalth caRe wOrkErS (HEROES) Study, Spain, 2021]

**Supplementary Table 1**.
Sampling strategy across health and care facilities in the main study locations (Andalucía, Madrid, and Murcia). [The COVID-19 HEalth caRe wOrkErS (HEROES) Study, Spain, 2021]

| Region | Recruitment unit | Type of unit | Sampling strategy |
|---|---|---|---|
| All | Sociedad Española de Medicina Preventiva, Salud Pública e Higiene (SEMPSPH) | Medical association | Representativeness approached by researcher; snowball sampling (text messages, emails) |
| All | Asociación Nacional de Psicólogos y Residentes (ANPIR) | Psychological association | Representativeness approached by researcher; snowball sampling (text messages, emails) |
| Cádiz | Hospital Universitario Puerto Real | General hospital | Representativeness approached by researcher |
| Granada | Hospital Regional Clínico Universitario San Cecilio | General hospital | Representativeness approached by researcher |
| Granada | Unidad de Gestión Clínica Almanjáyar | Outpatient specialty center | Representativeness approached by researcher |
| Granada | Unidad de Gestión Clínica Cartuja | Outpatient specialty center | Representativeness approached by researcher |
| Granada | Unidad de Gestión Clínica Gran Capitán | Outpatient specialty center | Representativeness approached by researcher |
| Huelva | Hospital Juan Ramón Jiménez | General hospital | Representativeness approached by researcher |
| Madrid | Colmenar Viejo Sur | Outpatient specialty center | Snowball sampling (text messages); billboard |
| Madrid | Hospital Universitario La Paz | General hospital | Pop-up alert embedded in electronic system; snowball sampling (text messages); billboard |
| Madrid | Oficina Regional de Salud Mental | Outpatient specialty center | Representativeness approached by researcher |
| Madrid | Hospital Universitario La Princesa | General hospital | Snowball sampling (text messages) |
| Madrid | Unión General de Trabajadores (UGT) | Labor union | Representativeness approached by researcher |
| Madrid | Comisiones Obreras (CCOO) | Labor union | Representativeness approached by researcher |
| Madrid | Sindicato de Enfermería (SATSE) | Labor union | Representativeness approached by researcher |
| Madrid | Asociación de Médicos y Titulados Superiores (AMYTS) | Labor union | Representativeness approached by researcher |
| Málaga | Unión General de Trabajadores (UGT) | Labor union | Representativeness approached by researcher |
| Málaga | Hospital Regional Universitario de Málaga | General hospital | Snowball sampling (text messages) |
| Málaga | Central Sindical Independiente y de Funcionarios (CSIF) | Labor union | Representativeness approached by researcher |
| Málaga | Comisiones Obreras (CCOO) | Labor union | Representativeness approached by researcher |
| Málaga | Hospital Universitario Virgen de la Victoria | General hospital | Snowball sampling (text messages) |
| Málaga | Sindicato de Enfermería (SATSE) | Labor union | Representativeness approached by researcher |

| | | | |
|---|---|---|---|
| Málaga | Sindicato de Técnicos Auxiliares de Enfermería (SAE) | Labor union | Representativeness approached by researcher |
| Málaga | Local non-hospital emergency departments (Málaga-Guadalhorce) | Non-hospital emergency department | Representativeness approached by researcher; snowball sampling (text messages, emails) |
| Málaga | Hospital Comarcal de la Axarquía | General hospital | Representativeness approached by researcher |
| Málaga | Hospital Comarcal Antequera | General hospital | Representativeness approached by researcher |
| Málaga | Medical Residents Coordination Office | Professional network | Representativeness approached by researcher |
| Málaga | Hospital San Juan de Dios | Private hospital | Representativeness approached by researcher |
| Málaga | Local health office (Norte de Málaga-Antequera) | Primary care center | Representativeness approached by researcher |
| Málaga | Local health office (Serranía de Málaga) | Primary care center | Representativeness approached by researcher |
| Málaga | Local health office (Este de Málaga-Axarquia) | Primary care center | Representativeness approached by researcher |
| Murcia | Hospital Clínico Universitario Virgen de la Arrixaca | General hospital | Representativeness approached by researcher; snowball sampling (text messages, emails) |
| Murcia | Hospital Los Arcos | General hospital | Representativeness approached by researcher; snowball sampling (text messages, emails) |
| Murcia | Oficina Regional de Salud Mental | Outpatient specialty center | Representativeness approached by researcher |
| Murcia | Instituto Murciano de Investigación Biomédica (IMIB) | Health research institute | Representativeness approached by researcher; snowball sampling (text messages, emails) |
| Murcia | Colegio de Médicos | Medical association | Representativeness approached by researcher |
| Murcia | Colegio de Enfermería | Nursing association | Representativeness approached by researcher |
| Murcia | Colegio de Farmacia | Pharmaceutical association | Representativeness approached by researcher |
| Murcia | Colegio de Fisioterapia | Physiotherapeutical association | Representativeness approached by researcher |
| Murcia | Comité Empresa (Sindicato) | Labor union | Representativeness approached by researcher |
| Murcia | Primary care network | Professional network | Representativeness approached by researcher |
| Sevilla | Hospital Universitario Virgen de Valme | General hospital | Representativeness approached by researcher |
| Sevilla | Área de Gestión Sanitaria Sur de Sevilla | Primary care center | Representativeness approached by researcher |

**Supplementary Table 2.**
Response rates across recruitment units. [The COVID-19 HEalth caRe wOrkErS (HEROES) Study, Spain, 2021]

| Region | Recruitment unit | Type of unit | Response rate (%) |
|---|---|---|---|
| Spain | Grupo 5 | Socio-community mental health centers | 28.2<br>67.6 [2] |
| Madrid | Hospital Universitario La Paz | General hospital | |
| | Emergency Department (Children's hospital) | | |
| | Nurses | | 3.1 |
| | Nurse technicians | | 7.7 |
| | Total | | 5.2 |
| | Emergency Department (Adult hospital) | | |
| | Nurses | | 6.7 |
| | Nurse technicians | | 3.5 |
| | Physicians | | 25.5 |
| | Total | | 8.6 |
| | Department of General Surgery (Adult hospital) | | |
| | Nurses | | 19.6 |
| | Nurse technicians | | 5.6 |
| | Physicians | | 13.4 |
| | Total | | 13.4 |
| | ICUs (Adult hospital) | | |
| | Nurses | | 10.9 |
| | Nurse technicians | | 2.7 |
| | Physicians | | 8.0 |
| | Total | | 8.1 |
| | ICU (Children's Hospital: nurses) | | 8.9 |
| | Department of Anesthesiology (physicians) | | 4.2 |
| | Department of Internal Medicine (physicians) | | 22.2 |
| | Department of Gastroenterology (physicians) | | 6.1 |
| | Department of Pneumology (physicians) | | 6.9 |
| | Department of Neurology (physicians) | | 7.7 |
| | Department of Mental Health | | |
| | Psychiatrists | | 34.1 |
| | Clinical psychologists | | 14.8 |
| | Total | | 26.8 |
| | Orderly team | | 26.1 |
| Andalucía | FAISEM | Socio-community mental health centers | 24.0 |
| Badajoz | Talarrubias | Primary care center | 90.0 |
| Granada | Almanjayar | Primary care center | 38.5 |
| Granada | Cartuja | Primary care center | 21.9 |
| Granada | Gran Capitán | Primary care center | 12.3 |
| Málaga | Alameda Perchel | Primary care center | 36.8 |
| Málaga | Rincón de la Victoria | Primary care center | 8.3 |
| Málaga | Urgencias Distrito Hospital Clínico | Emergency Unit | 14.6 |
| Murcia | Los Barreros | Primary care center | 68.2 |
| Murcia | Isaac Peral | Primary care center | 27.5 |
| Murcia | San Andrés | Primary care center | 16.7 |
| Murcia | Mental health center of Cartagena | Mental health center | 80.4 |
| Murcia | Consejería de Salud (COVID-19 contact tracers) | Regional Health Office | 81.3 |
| Murcia | Consejería de Salud (others) | Regional Health Office | 35.0 |
| Murcia | Hospital Clínico Universitario Virgen de la Arrixaca | General hospital | |
| | Department of Pediatrics | | 100 |
| | Department of Preventive Medicine | | 94.4 |
| | Department of Oncology | | 5.7 |

**Supplementary Table 3**.

Psychological distress, depression symptoms, and posttraumatic stress disorder symptoms among all follow-up respondents (N = 1,807). [The COVID-19 HEalth caRe wOrkErS (HEROES) Study, Spain, 2021]

| | Psychological distress (GHQ-12) | | Depression symptoms (PHQ-9) | | PTSD symptoms (PC-PTSD-5) | |
|---|---|---|---|---|---|---|
| | n (%) [a] | M (SD) [b] | n (%) [a] | M (SD) [b] | n (%) [a] | M (SD) [b] |
| Overall | 881 (56) | 3.8 (3.4) | 326 (21) | 6.3 (5.1) | 781 (51) | 2.6 (1.6) |
| Age group | | | | | | |
| 18-35 | 290 (59) | 4 (3.3) | 115 (25) | 6.9 (4.8) | 273 (60) | 2.9 (1.5) |
| 36-50 | 367 (56) | 3.9 (3.5) | 140 (22) | 6.5 (5.2) | 334 (53) | 2.6 (1.6) |
| Over 50 | 204 (52) | 3.6 (3.5) | 64 (16) | 5.6 (5.1) | 157 (40) | 2.2 (1.6) |
| Gender | | | | | | |
| Male | 166 (48) | 3.3 (3.5) | 45 (13) | 5.3 (5.1) | 133 (40) | 2.1 (1.5) |
| Female | 714 (58) | 3.9 (3.4) | 281 (23) | 6.7 (5) | 648 (55) | 2.7 (1.6) |
| Educational level | | | | | | |
| Primary studies | 5 (36) | 3 (3.7) | 2 (13) | 5 (6.6) | 6 (40) | 2.1 (1.7) |
| Secondary studies | 176 (51) | 3.7 (3.5) | 75 (22) | 6.6 (5.5) | 181 (54) | 2.7 (1.6) |
| University studies | 698 (57) | 3.9 (3.4) | 249 (21) | 6.3 (4.9) | 591 (51) | 2.5 (1.6) |
| Parental educational level | | | | | | |
| Primary studies | 267 (52) | 3.6 (3.4) | 96 (19) | 6 (5) | 245 (49) | 2.5 (1.6) |
| Secondary studies | 275 (58) | 3.9 (3.3) | 96 (20) | 6.6 (4.8) | 250 (55) | 2.7 (1.5) |
| University studies | 306 (56) | 3.9 (3.5) | 121 (23) | 6.3 (5.1) | 258 (50) | 2.5 (1.6) |
| Type of job | | | | | | |
| Physicians | 233 (60) | 4.2 (3.5) | 82 (21) | 6.5 (5.1) | 183 (48) | 2.4 (1.7) |
| Nurses | 172 (58) | 4.1 (3.6) | 75 (27) | 7 (5.1) | 174 (64) | 3 (1.5) |
| Health technicians [c] | 44 (55) | 4.1 (3.8) | 24 (32) | 8.1 (6.2) | 48 (67) | 3 (1.6) |
| Other HCWs [d] | 131 (52) | 3.2 (2.9) | 42 (17) | 5.6 (4.4) | 117 (47) | 2.4 (1.6) |
| Ancillary workers [e] | 74 (51) | 3.5 (3.5) | 26 (18) | 6.1 (5.3) | 64 (45) | 2.5 (1.6) |
| Residential support workers | 179 (51) | 3.6 (3.4) | 60 (17) | 5.9 (4.9) | 163 (49) | 2.5 (1.5) |
| Other | 48 (63) | 4.1 (3.2) | 17 (22) | 6.7 (5.1) | 32 (44) | 2.3 (1.3) |

*Note*

All percentages are valid percentages

GHQ-12 = General Health Questionnaire – 12, PHQ-9 = Patient Health Questionnaire – 9, PTSD = posttraumatic stress disorder, PC-PTSD-5 = Primary Care PTSD Screen for the DSM-5, HCWs = healthcare workers, PPE = personal protective equipment

[a] Number of respondents screening positive for mental health problems (cutoffs: PHQ-9 > 9, GHQ-12 > 2, and PC-PTSD > 2)

[b] Means and standard deviations of the total scores of the PHQ-9 (range: 0-27), the GHQ-12 (range: 0-12), and the PC-PTSD-5 (range: 0-5)

[c] Health technicians include nurse, X-ray, or laboratory technicians, among others

[d] Other HCWs include clinical psychologists, physiotherapists, or biologists, among others

[e] Ancillary workers include security staff, drivers, administrative staff, or cleaning staff, among others

**Supplementary Table 4**.

Association between participants' sociodemographic characteristics and COVID-19-related exposures, measured at baseline, and the probability of screening positive for psychological distress, depressive symptoms, and posttraumatic stress disorder symptoms, at follow-up (8 months). [The COVID-19 HEalth caRe wOrkErS (HEROES) Study, Spain, 2021]

| | Psychological distress (GHQ-12) | | | | Depression symptoms (PHQ-9) | | | | PTSD symptoms (PC-PTSD-5) | | | |
|---|---|---|---|---|---|---|---|---|---|---|---|---|
| | Unadjusted | | Adjusted | | Unadjusted | | Adjusted | | Unadjusted | | Adjusted | |
| | OR | 95 percent CI | OR | 95 percent CI | OR | 95 percent CI | OR | 95 percent CI | OR | 95 percent CI | OR | 95 percent CI |
| 18-35 years old [ref: > 50] [a] | 1.03 | (0.67, 1.58) | 0.94 | (0.61, 1.46) | 1.60 | (0.94, 2.71) | 1.35 | (0.78, 2.33) | 1.77 | (1.15, 2.74) | 1.58 | (1.01, 2.46) |
| 35-50 years old [ref: > 50] [a] | 0.89 | (0.6, 1.33) | 0.84 | (0.56, 1.26) | 1.24 | (0.75, 2.07) | 1.15 | (0.69, 1.93) | 1.31 | (0.87, 1.96) | 1.23 | (0.82, 1.86) |
| Female gender [ref: male] [b] | 1.82 | (1.2, 2.77) | 1.77 | (1.15, 2.72) | 3.62 | (1.75, 7.46) | 3.21 | (1.54, 6.69) | 1.91 | (1.24, 2.93) | 1.73 | (1.11, 2.7) |
| University studies [ref: primary/secondary] [c] | 0.76 | (0.46, 1.23) | 0.76 | (0.44, 1.3) | 0.61 | (0.36, 1.03) | 0.58 | (0.32, 1.05) | 0.78 | (0.48, 1.27) | 0.78 | (0.45, 1.33) |
| Frontline position [ref: no] [c] | 0.92 | (0.67, 1.28) | 0.87 | (0.61, 1.24) | 0.92 | (0.61, 1.38) | 0.86 | (0.54, 1.36) | 1.68 | (1.21, 2.35) | 1.75 | (1.2, 2.54) |
| Adequate access to PPE [ref: inadequate] [c] | 0.61 | (0.44, 0.85) | 0.66 | (0.47, 0.93) | 0.72 | (0.48, 1.09) | 0.83 | (0.54, 1.29) | 0.58 | (0.41, 0.81) | 0.63 | (0.44, 0.9) |
| Fear of getting infected [ref: none or low] [c] | 1.32 | (0.94, 1.86) | 1.29 | (0.9, 1.84) | 1.19 | (0.77, 1.83) | 1.10 | (0.69, 1.74) | 1.79 | (1.26, 2.53) | 1.69 | (1.16, 2.45) |
| Fear of infecting loved ones [ref: none or low] [c] | 1.15 | (0.77, 1.72) | 1.20 | (0.78, 1.85) | 0.98 | (0.59, 1.62) | 0.88 | (0.51, 1.52) | 1.96 | (1.3, 2.97) | 2.16 | (1.37, 3.4) |

*Note*

GHQ-12 = General Health Questionnaire – 12 items, PHQ-9 = Patient Health Questionnaire – 9 items, PC-PTSD-5 = Primary Care PTSD Screen for DSM-5, OR = odds ratio, CI = confidence interval, PPE = personal protective equipment

[a] Adjusted for gender

[b] Adjusted for age

[c] Adjusted for age, gender, and region-level 14-day COVID-19 cumulative incidence (fixed factor)

**Supplementary Table 5.**

Association between participants' sociodemographic characteristics and COVID-19-related exposures, measured at baseline, and mental health outcomes' total scores (psychological distress, depressive symptoms, and posttraumatic stress disorder symptoms), measured at follow-up (8 months). [The COVID-19 HEalth caRe wOrkErS (HEROES) Study, Spain, 2021]

| | Psychological distress (GHQ-12) | | | | Depression symptoms (PHQ-9) | | | | PTSD symptoms (PC-PTSD-5)* | | | |
|---|---|---|---|---|---|---|---|---|---|---|---|---|
| | Unadjusted | | Adjusted | | Unadjusted | | Adjusted | | Unadjusted | | Adjusted | |
| | B | 95 percent CI | B | 95 percent CI | B | 95 percent CI | B | 95 percent CI | B | 95 percent CI | B | 95 percent CI |
| Age in years [a] | 0.00 | (-0.03, 0.02) | 0.01 | (-0.01, 0.04) | -0.04 | (-0.08, 0) | 0.01 | (-0.02, 0.04) | -0.02 | (-0.04, -0.01) | -0.02 | (-0.03, -0.01) |
| Female gender [b] | 1.05 | (0.32, 1.77) | 0.32 | (-0.37, 1.01) | 2.14 | (1.1, 3.18) | 0.57 | (-0.32, 1.46) | 0.68 | (0.34, 1.02) | 0.31 | (-0.02, 0.64) |
| Educational level [c] | -0.22 | (-0.57, 0.12) | -0.29 | (-0.63, 0.05) | -0.54 | (-1.02, -0.05) | -0.36 | (-0.79, 0.07) | -0.16 | (-0.32, 0) | -0.23 | (-0.38, -0.07) |
| Frontline position [c] | 0.13 | (-0.42, 0.69) | -0.31 | (-0.88, 0.25) | 0.08 | (-0.72, 0.89) | -0.68 | (-1.41, 0.04) | 0.46 | (0.2, 0.73) | 0.33 | (0.06, 0.6) |
| Adequate access to PPE [c] | -0.53 | (-0.86, -0.2) | -0.20 | (-0.52, 0.12) | -1.03 | (-1.5, -0.56) | -0.41 | (-0.81, 0) | -0.37 | (-0.52, -0.21) | -0.21 | (-0.36, -0.06) |
| Fear of getting infected [c] | 0.25 | (-0.13, 0.63) | -0.20 | (-0.58, 0.18) | 0.42 | (-0.12, 0.96) | -0.30 | (-0.78, 0.17) | 0.46 | (0.28, 0.64) | 0.27 | (0.09, 0.45) |
| Fear of infecting loved ones [c] | 0.32 | (-0.02, 0.66) | 0.13 | (-0.2, 0.46) | 0.66 | (0.17, 1.16) | 0.03 | (-0.4, 0.45) | 0.50 | (0.34, 0.65) | 0.42 | (0.27, 0.57) |

*Note*

GHQ-12 = General Health Questionnaire – 12 items, PHQ-9 = Patient Health Questionnaire – 9 items, PC-PTSD-5 = Primary Care PTSD Screen for DSM-5, B = beta, CI = confidence interval, PPE = personal protective equipment

[a] Adjusted for gender and GHQ-12 / PHQ-9 baseline total score

[b] Adjusted for age and GHQ-12 / PHQ-9 baseline total score

[c] Adjusted for age, gender, GHQ-12 / PHQ-9 baseline total score, and region-level 14-day COVID-19 cumulative incidence (fixed factor)

* Baseline assessment did not include PC-PTSD-5. Models are corrected using GHQ-12 baseline total score instead

**Supplementary Table 6**.

Association between participants' sociodemographic characteristics and COVID-19-related exposures, measured at baseline, and the probability of screening positive for psychological distress, depressive symptoms, and posttraumatic stress disorder symptoms, at follow-up (8 months). [The COVID-19 HEalth caRe wOrkErS (HEROES) Study, Spain, 2021]

| | Psychological distress (GHQ-12) | | | | Depression symptoms (PHQ-9) | | | | PTSD symptoms (PC-PTSD-5)* | | | |
|---|---|---|---|---|---|---|---|---|---|---|---|---|
| | Unadjusted | | Adjusted | | Unadjusted | | Adjusted | | Unadjusted | | Adjusted | |
| | OR | 95 percent CI | OR | 95 percent CI | OR | 95 percent CI | OR | 95 percent CI | OR | 95 percent CI | OR | 95 percent CI |
| 18-35 years old [ref: > 50] [a] | 1.03 | (0.67, 1.58) | 0.92 | (0.57, 1.48) | 1.60 | (0.94, 2.71) | 1.14 | (0.61, 2.12) | 1.77 | (1.15, 2.74) | 1.59 | (0.97, 2.61) |
| 35-50 years old [ref: > 50] [a] | 0.89 | (0.6, 1.33) | 0.77 | (0.49, 1.2) | 1.24 | (0.75, 2.07) | 1.02 | (0.56, 1.84) | 1.31 | (0.87, 1.96) | 1.21 | (0.76, 1.9) |
| Female [ref: male] [b] | 1.82 | (1.2, 2.77) | 1.36 | (0.85, 2.16) | 3.62 | (1.75, 7.46) | 2.34 | (1.08, 5.08) | 1.91 | (1.24, 2.93) | 1.33 | (0.82, 2.16) |
| University studies [ref: primary/secondary] [c] | 0.76 | (0.46, 1.23) | 0.69 | (0.37, 1.26) | 0.61 | (0.36, 1.03) | 0.70 | (0.35, 1.42) | 0.78 | (0.48, 1.27) | 0.79 | (0.43, 1.44) |
| Frontline position [ref: no] [c] | 0.92 | (0.67, 1.28) | 0.82 | (0.55, 1.2) | 0.92 | (0.61, 1.38) | 0.78 | (0.46, 1.31) | 1.68 | (1.21, 2.35) | 1.71 | (1.14, 2.58) |
| Adequate access to PPE [ref: inadequate] [c] | 0.61 | (0.44, 0.85) | 0.71 | (0.5, 1.03) | 0.72 | (0.48, 1.09) | 0.84 | (0.51, 1.38) | 0.58 | (0.41, 0.81) | 0.71 | (0.48, 1.05) |
| Fear of getting infected [ref: none or low] [c] | 1.32 | (0.94, 1.86) | 1.05 | (0.71, 1.54) | 1.19 | (0.77, 1.83) | 1.00 | (0.6, 1.66) | 1.79 | (1.26, 2.53) | 1.18 | (0.78, 1.79) |
| Fear of infecting loved ones [ref: none or low] [c] | 1.15 | (0.77, 1.72) | 1.05 | (0.66, 1.68) | 0.98 | (0.59, 1.62) | 0.64 | (0.35, 1.2) | 1.96 | (1.3, 2.97) | 1.99 | (1.2, 3.29) |

*Note*.

GHQ-12 = General Health Questionnaire – 12 items, PHQ-9 = Patient Health Questionnaire – 9 items, PC-PTSD-5 = Primary Care PTSD Screen for DSM-5, OR = odds ratio, CI = confidence interval, PPE = personal protective equipment

[a] Adjusted for gender and probable psychological distress (GHQ-12) or depression (PHQ-9) at baseline

[b] Adjusted for age and probable psychological distress (GHQ-12) or depression (PHQ-9) at baseline

[c] Adjusted for age, gender, probable psychological distress (GHQ-12) or depression (PHQ-9) at baseline, and region-level 14-day COVID-19 cumulative incidence (fixed factor)

* Baseline assessment did not include PC-PTSD-5. Models are corrected using GHQ-12 baseline total score instead

Cut-off scores used: GHQ-12 > 2, PHQ-9 > 9, and PC-PTSD-5 > 2
